# Demographic and economic characteristics of residents of four subsidized housing facilities in Nashville, TN, and their preferences for co-located services

**DOI:** 10.1101/2025.08.23.25334285

**Authors:** Kate Clouse, Christine Kimpel, Lorely Chávez, Terry Terrell, Julia Landivar Donato, Natalie Hasbrooke, Makenna Frenia, Brent Elrod, Christian Ketel

## Abstract

**Introduction:** Subsidized housing provides affordable rent to low-income households, many of whom have high rates of disability and chronic disease, but it is generally not designed to deliver the health and social services these residents may need. In this study, we sought to characterize residents of four subsidized housing facilities in Nashville, Tennessee, an area marked by rapid growth, poverty, and housing instability, and to assess their preferences for co-located services.

**Methods:** A cross-sectional study (2023-2025) recruited 140 residents from four subsidized housing facilities using randomized selection. We administered semi-structured questionnaires assessing demographics, past experiences with homelessness and incarceration, health, and preferences for health and social services.

**Results:** Most residents were Black/African American (63.6%) and older adults, with 80% earning ≤c$20,000 annually. Most had experienced homelessness (70.0%) and over half incarceration (53.6%). Residents expressed strong interest in on-site services, particularly dental and vision care (over 90%) and adult primary care accepting TennCare/Medicaid (91.4%).

**Discussion:** Findings depict a population facing multiple vulnerabilities, including economic instability, social isolation, and histories of homelessness and incarceration. This study highlights the importance of combining housing with comprehensive health and social services in addressing inequities and prioritizing resident feedback in service design.

## Introduction

The built environment, including housing, is an essential determinant of health.^1^ Subsidized housing, including public housing, project-based rental assistance, and housing choice vouchers, gives low-income individuals and families access to stable, affordable homes and can be an important pathway out of homelessness.^2^ However, residents of subsidized housing bear a disproportionate burden of chronic disease and disability, even compared with other low-income renters.^3–5^ Despite these needs, most subsidized housing is not designed to deliver health or social services, and federally funded service coordination is concentrated in properties for older adults and people with disabilities.^6^

While research on subsidized and transitional housing has expanded in recent years, most studies have focused on populations in the Northeast United States.^7–11^ Less is known about resident characteristics and service needs in the Southern U.S., where poverty and housing instability are disproportionately high. The South carries the nation’s highest poverty burden,^12^ and Tennessee has experienced a recent increase in poverty, compounding barriers to housing security.^13^ Nashville, the state capital and third-largest city in the Southeast, is marked by rapid growth, wealth inequality, and a chronic undersupply of affordable housing.^14,15^ Nearly half of the city’s 300,000 renters are considered cost-burdened, paying more than 30% of their income toward rent.^16^

Effective housing interventions must be grounded in understanding residents’ specific needs and lived experiences, particularly in underrepresented regions. Studies have shown that housing stability alone does not guarantee improved health or psychosocial outcomes. Community engagement in the design and evaluation of housing services may enhance feasibility, acceptability, and alignment with residents’ priorities.^17^

To address this gap, Vanderbilt University School of Nursing researchers partnered with Urban Housing Solutions (UHS), Nashville’s largest private nonprofit affordable housing provider.^18^ UHS follows a resident services model incorporating resident-centered services and collaborative programming to promote long-term housing retention and wellness. This study aimed to describe residents’ demographic and economic characteristics across four subsidized housing facilities in Nashville and assess their preferences for co-located health and social services to inform future program design and address persistent inequities in access to supportive housing services in the U.S. South by engaging directly with residents.

## Methods

### Study Sites

During this cross-sectional study, we enrolled participants at four subsidized housing facilities owned by UHS. At each facility, we received a list of occupied units from the building manager; these anonymized units were randomly ordered using the uniform random number generator in SPSS version 29.0. We then sequentially recruited individuals based on this list. To ensure balanced sampling across varying-sized facilities, we targeted approximately 35-40% of occupied units per site. One facility (Site D) offers subsidized and market-rate units designated for older adults (aged 62+); only subsidized units were included in the sampling frame.

UHS supports residents through multiple housing subsidy programs. These include Section 8 Project-Based Rental Assistance (PBRA) and Housing Choice Vouchers, both federally funded and locally administered by the Metropolitan Development and Housing Agency (MDHA), which cap tenant rent contributions at roughly 30% of household income.^19,20^ Many UHS properties are also supported through the Low-Income Housing Tax Credit (LIHTC) program for households earning 30-60% of the area median income.^21^ Additionally, some residents receive support via the Housing Opportunities for Persons with AIDS (HOPWA) program, which provides housing and supportive services for individuals living with HIV/AIDS.^22^

### Recruitment and enrollment

From September 2023 to June 2025, two study team members knocked on the door of the listed units for recruitment. If the resident was home and completed the screening consent, they introduced the study and asked if the resident was interested in participating, scheduling accordingly. If the resident was not home, an invitation letter printed in English and Spanish was taped to their door with contact information for follow-up. Each apartment door was approached four times before determining that the resident was not interested. If more than one resident lived in the unit, we invited the person whose name was on the lease. Recruitment efforts were aided by advertising flyers posted in common areas throughout the facility. All study materials were available in English and Spanish, and members of our study team were bilingual in both languages. All study materials were completed directly into REDCap using a tablet computer. REDCap is a secure, encrypted, web-based software platform that supports data capture for research studies.^23^ At the end of the study, data were exported from REDCap as .csv files for analysis.

Individuals were eligible for study participation if they were adults (age 18+) living in the selected UHS unit. Participants were excluded if they were unable to provide informed consent or respond to the questionnaire due to cognitive, auditory, or visual impairment or a language barrier (language other than English or Spanish).

### Study procedures

All study activities were conducted in a single visit. A study coordinator administered a semi-structured questionnaire after providing electronic informed consent, including assessing understanding using teach-back methods. The questionnaire lasted about 45 minutes and included questions related to demographics, previous experience with homelessness or incarceration (defined as more than two nights in a row in jail, prison, or a detention center or juvenile correctional facility), housing, transportation and health. We assessed preferences for future services by asking, “I’d like to ask you to imagine a center based at this property that could offer care and services to the residents. For each of the following statements, I’d like to know how you feel using a scale of ‘not at all interested,’ ‘somewhat interested,’ or ‘very interested.’” We combined “somewhat interested” and “very interested” in the analysis.

After completion of the questionnaire, participants were remunerated for their time with a physical $50 gift card to a major supermarket chain with numerous locations throughout Nashville. Based on the low email use in this population noted in preliminary work, we received special permission to distribute physical gift cards, rather than the standard electronic gift cards used for other studies at our institution.

### Statistical analysis

Categorical data are presented as counts and proportions; continuous data are presented as medians and interquartile ranges (IQR). We obtained p-values, with α=0.05, by comparing proportions using the Chi-square test, or Fisher exact test for small cell sizes (n≤c5) and compared medians using the Kruskal-Wallis test. Data were analyzed using SAS, version 9.4 (SAS Institute, Cary, NC).

### Ethical consideration

All participants provided electronic informed consent prior to participation, and a hard copy of the consent was given to each participant. Study activities and materials were approved by the Institutional Review Board of Vanderbilt University (approval number 230916).

## Results

### Participant characteristics

We approached 285 units; at 143 of these, the residents refused (n=65), could not be reached (n=58), failed to show to an appointment (n=18), or came for an appointment after our recruitment goal was met (n=2). Two participants were withdrawn for declining to complete the questionnaire once consented. Overall, we enrolled 140 participants, representing 36.6% of possible units, with a range of 35.1-39.2% (p=0.97) across the four facilities. Median (IQR) household size was 1 (1-1) person; at Site C, 35% reported more than one person (range 2-7) in the household. Participant characteristics are summarized in **Table 1**. More males participated overall (55.7%), and this ranged from 41.8% at Site B to 82.5% at Site A. Median (IQR) age was 61.5 (50-67) years; as expected, Site D had the greatest proportion of participants (84.0%) in the age 65+ category.

**Table 1.** Characteristics of participants in a study of residents living in subsidized housing in Nashville, TN, 2023-2025 (n=140).

|  | **Overall** | **Site A** | **Site B** | **Site C** | **Site D** | **p-value** |
|---|---|---|---|---|---|---|
| Participants/Available units (%) | 140/383 (36.6%) | 40/102 (39.2%) | 55/156 (35.2%) | 20/57 (35.1%) | 25/68 (36.8%) | 0.971 |
| Sex |  |  |  |  |  |  |
| Male | 78 (55.7) | 33 (82.5) | 23 (41.8) | 9 (45.0) | 13 (52.0) | 0.0007 |
| Female | 62 (44.3) | 7 (17.5) | 32 (58.2) | 11 (55.0) | 12 (48.0) |  |
| Age (years), median (IQR) | 61.5 (50-67) | 63 (52-67) | 57 (45-63) | 52 (41-62) | 70 (66-73) | <0.0001 |
| Age (years) |  |  |  |  |  |  |
| 18-39 | 16 (11.4) | 1 (2.5) | 11 (20.0) | 4 (20.0) | 0 (0.0) | <0.0001 |
| 40-64 | 77 (55.0) | 24 (60.0) | 37 (67.3) | 12 (60.0) | 4 (16.0) |  |
| 65+ | 47 (33.6) | 15 (37.5) | 7 (12.7) | 4 (20.0) | 21 (84.0) |  |
| Race (self-identified)* |  |  |  |  |  |  |
| Black/African American | 89 (63.6) | 20 (50.0) | 35 (63.6) | 13 (65.0) | 21 (84.0) | <0.0001 |
| White | 41 (29.3) | 13 (32.5) | 19 (34.6) | 5 (25.0) | 4 (16.0) |  |
| American Indian/Alaska Native | 6 (4.3) | 5 (12.5) | 1 (1.8) | 0 (0.0) | 0 (0.0) |  |
| Other | 2 (1.4) | 2 (5.0) | 0 (0.0) | 0 (0.0) | 0 (0.0) |  |
| Prefer not to say | 2 (1.4) | 0 (0.0) | 0 (0.0) | 2 (10.0) | 0 (0.0) |  |
| Identify as Hispanic or Latino** |  |  |  |  |  |  |
| Yes | 7 (5.0) | 3 (7.5) | 0 (0.0) | 4 (20.0) | 0 (0.0) | 0.0003 |
| No | 132 (95.0) | 37 (92.5) | 54 (100.0) | 16 (80.0) | 25 (100.0) |  |
| Nationality |  |  |  |  |  |  |
| Born in the US | 132 (95.0) | 40 (100.0) | 53 (96.4) | 16 (80.0) | 24 (96.0) | 0.001 |
| Born outside of the US | 7 (5.0) | 0 (0.0) | 2 (3.6) | 4 (20.0) | 1 (4.0) |  |
| Primary language** |  |  |  |  |  |  |
| English | 135 (97.1) | 40 (100.0) | 53 (98.2) | 17 (85.0) | 25 (100.0) | 0.001 |
| Spanish | 3 (2.2) | 0 (0.0) | 0 (0.0) | 3 (15.0) | 0 (0.0) |  |
| Arabic | 1 (0.7) | 0 (0.0) | 1 (1.9) | 0 (0.0) | 0 (0.0) |  |
| Currently unmarried or not partnered |  |  |  |  |  |  |
| Yes | 136 (97.1) | 3 (7.5) | 1 (1.8) | 0 (0.0) | 0 (0.0) | 0.035 |
| No | 4 (2.9) | 37 (92.5) | 54 (98.2) | 20 (100.0) | 25 (100.0) |  |
| Sexual orientation |  |  |  |  |  |  |
| Straight | 121 (86.4) | 36 (90.0) | 45 (81.8) | 18 (90.0) | 22 (88.0) | <0.0001 |
| Gay or lesbian | 8 (5.7) | 2 (5.0) | 4 (7.3) | 0 (0.0) | 2 (8.0) |  |
| Bisexual | 7 (5.0) | 1 (2.5) | 5 (9.1) | 0 (0.0) | 1 (4.0) |  |
| None of these | 4 (2.9) | 1 (2.5) | 1 (1.8) | 2 (10.0) | 0 (0.0) |  |
| Education |  |  |  |  |  |  |
| Less than high school | 35 (25.0) | 12 (30.0) | 12 (21.8) | 7 (35.0) | 4 (16.0) | <0.0001 |
| Completed high school | 55 (39.3) | 15 (37.5) | 25 (45.5) | 5 (25.0) | 10 (40.0) |  |
| More than high school | 50 (35.7) | 13 (32.5) | 18 (32.7) | 8 (40.0) | 11 (44.0) |  |
| Prior military service** |  |  |  |  |  |  |
| Yes | 15 (10.8) | 7 (17.5) | 1 (1.9) | 3 (15.0) | 4 (16.0) | 0.0003 |
| No | 124 (89.2) | 33 (88.2) | 53 (98.2) | 17 (85.0) | 21 (84.0) |  |

\* Participants could select more than one race; results sum to more than 100%
\*\* Removed one participant (for each category) who preferred not to answer question or was not sure

Most participants self-identified as Black/African-American race (63.6%), and this ranged from 50-84% across sites. Overall, few participants (5%) identified as Hispanic/Latino, and most were born in the US (95%) and spoke English as their primary language (97.1%). One exception is at Site C, where 4/20 (20%) of participants identified as Hispanic/Latino and were born outside of the US. Of our 140 participants, seven participants were born outside of the US, including Germany, Honduras, Jamaica, Mexico and Sudan.

Only four participants overall (2.9%) reported being currently married or partnered and 13.4% selected a sexual identity other than “straight.” Fewer than two-thirds (64.3%) of respondents indicated they had completed high school. Overall, 10.8% of participants reported previous military service, but this varied significantly from 1.9% at Site B to 17.5% at Site A (p=0.0003).

### Economic indicators

**Table 2** displays key economic indicators of our sample. Many participants (37.9%) were unemployed, but not seeking work; this ranged from 20% at Site C to 43.6% at Site B. More than one-quarter (27.1%) were retired, including 60% of Site D residents. Site C represented the highest proportion of participants employed at 35%, and underemployed, with 30% working part-time or seeking work.

**Table 2.**
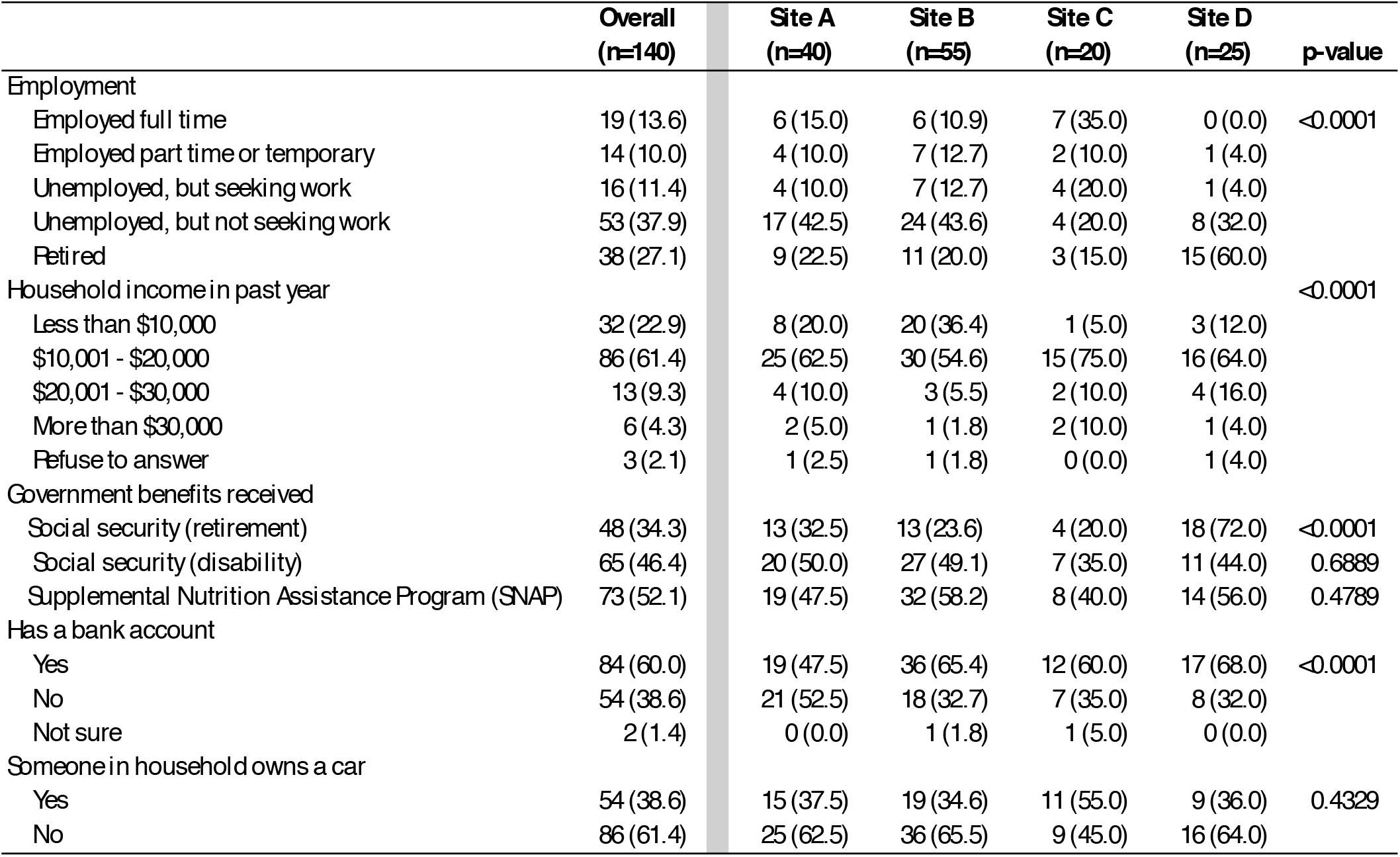
Economic indicators among 140 residents of subsidized housing facilities in Nashville, TN, 2023-2025.

|  | Overall<br>(n=140) | Site A<br>(n=40) | Site B<br>(n=55) | Site C<br>(n=20) | Site D<br>(n=25) | p-value |
|---|---|---|---|---|---|---|
| Employment |  |  |  |  |  |  |
| Employed full time | 19 (13.6) | 6 (15.0) | 6 (10.9) | 7 (35.0) | 0 (0.0) | <0.0001 |
| Employed part time or temporary | 14 (10.0) | 4 (10.0) | 7 (12.7) | 2 (10.0) | 1 (4.0) |  |
| Unemployed, but seeking work | 16 (11.4) | 4 (10.0) | 7 (12.7) | 4 (20.0) | 1 (4.0) |  |
| Unemployed, but not seeking work | 53 (37.9) | 17 (42.5) | 24 (43.6) | 4 (20.0) | 8 (32.0) |  |
| Retired | 38 (27.1) | 9 (22.5) | 11 (20.0) | 3 (15.0) | 15 (60.0) |  |
| Household income in past year |  |  |  |  |  | <0.0001 |
| Less than \$10,000 | 32 (22.9) | 8 (20.0) | 20 (36.4) | 1 (5.0) | 3 (12.0) |  |
| \$10,001 - \$20,000 | 86 (61.4) | 25 (62.5) | 30 (54.6) | 15 (75.0) | 16 (64.0) |  |
| \$20,001 - \$30,000 | 13 (9.3) | 4 (10.0) | 3 (5.5) | 2 (10.0) | 4 (16.0) |  |
| More than \$30,000 | 6 (4.3) | 2 (5.0) | 1 (1.8) | 2 (10.0) | 1 (4.0) |  |
| Refuse to answer | 3 (2.1) | 1 (2.5) | 1 (1.8) | 0 (0.0) | 1 (4.0) |  |
| Government benefits received |  |  |  |  |  |  |
| Social security (retirement) | 48 (34.3) | 13 (32.5) | 13 (23.6) | 4 (20.0) | 18 (72.0) | <0.0001 |
| Social security (disability) | 65 (46.4) | 20 (50.0) | 27 (49.1) | 7 (35.0) | 11 (44.0) | 0.6889 |
| Supplemental Nutrition Assistance Program (SNAP) | 73 (52.1) | 19 (47.5) | 32 (58.2) | 8 (40.0) | 14 (56.0) | 0.4789 |
| Has a bank account |  |  |  |  |  |  |
| Yes | 84 (60.0) | 19 (47.5) | 36 (65.4) | 12 (60.0) | 17 (68.0) | <0.0001 |
| No | 54 (38.6) | 21 (52.5) | 18 (32.7) | 7 (35.0) | 8 (32.0) |  |
| Not sure | 2 (1.4) | 0 (0.0) | 1 (1.8) | 1 (5.0) | 0 (0.0) |  |
| Someone in household owns a car |  |  |  |  |  |  |
| Yes | 54 (38.6) | 15 (37.5) | 19 (34.6) | 11 (55.0) | 9 (36.0) | 0.4329 |
| No | 86 (61.4) | 25 (62.5) | 36 (65.5) | 9 (45.0) | 16 (64.0) |  |

At each site, most participants reported annual household income in the $10,001-20,000 category, representing 61.4% overall. Site B had the highest proportion of participants (36.4%) in the lowest income category of less than $10,000. Overall, only six participants (4.3%) reported household income above $30,000. Enrollment in government benefit programs was widely reported in our sample, including Social Security retirement (34.3%), Social Security disability (46.4%), and Supplemental Nutrition Assistance Program (SNAP) (52.1%). Government program participation was lowest at Site C, where income and employment were highest.

Overall, more than one-third (38.6%) reported having no bank account; Site A was highest at 52.5%. Nearly two-thirds (61.4%) overall reported that no one in the household owned a car; this was lowest (45%) at Site C.

### Homelessness and incarceration

**Table 3** displays results of experience with homelessness and incarceration. Overall, 70.0% of participants reported previously experiencing homelessness, ranging from 48% at Site D to 78.2% at Site B. Most (75.5%) reported experiences of homelessness occurred more than three years ago. The majority (71.4%) reported experiencing homelessness lasting more than one year. Among this group, the median (IQR) duration of homelessness was 5 (2-10) years, with Site A reporting the longest duration at 7 (3-15) years.

**Table 3.** Experience with homelessness and incarceration among 140 residents of subsidized housing facilities in Nashville, TN, 2023-2025.

|  | Overall<br>(n=140) | Site A<br>(n=40) | Site B<br>(n=55) | Site C<br>(n=20) | Site D<br>(n=25) | p-value |
|---|---|---|---|---|---|---|
| **Homelessness** |  |  |  |  |  |  |
| Ever experienced |  |  |  |  |  | 0.0229 |
| Yes | 98 (70.0) | 31 (77.5) | 43 (78.2) | 12 (60.0) | 12 (48.0) |  |
| No | 42 (30.0) | 9 (22.5) | 12 (21.8) | 8 (40.0) | 13 (52.0) |  |
| Timing of homelessness (n=98) |  |  |  |  |  |  |
| Within the past year | 15 (15.3) | 4 (12.9) | 8 (18.6) | 1 (8.3) | 2 (16.7) | 0.0001 |
| More than 1 year ago, but less than 3 years | 9 (9.2) | 1 (3.2) | 7 (16.3) | 1 (8.3) | 0 (0.0) |  |
| More than 3 years ago | 74 (75.5) | 26 (83.9) | 28 (65.1) | 10 (83.3) | 10 (83.3) |  |
| Duration of homelessness (n=98) |  |  |  |  |  |  |
| More than 1 year | 70 (71.4) | 23 (74.2) | 32 (74.4) | 6 (50.0) | 9 (75.0) | 0.0036 |
| Less than 1 year | 28 (28.6) | 8 (25.8) | 11 (25.6) | 6 (50.0) | 3 (25.0) |  |
| Median time in years (IQR), if more than one year (n=70) | 5 (2-10) | 7 (3-15) | 3 (2-7) | 3 (3-5) | 5 (4-7) | 0.1542 |
| **Incarceration** |  |  |  |  |  |  |
| Ever experienced |  |  |  |  |  | 0.0300 |
| Yes | 75 (53.6) | 28 (70.0) | 30 (54.6) | 8 (40.0) | 9 (36.0) |  |
| No | 65 (46.4) | 12 (30.0) | 25 (45.5) | 12 (60.0) | 16 (64.0) |  |
| Timing of incarceration (n=75) |  |  |  |  |  | 0.0144 |
| Within the past year | 7 (9.3) | 1 (3.6) | 3 (10.0) | 2 (25.0) | 1 (11.1) |  |
| More than 1 year ago | 68 (90.7) | 27 (96.4) | 27 (90.0) | 6 (75.0) | 8 (88.9) |  |
| Duration of incarceration (n=74)* |  |  |  |  |  | 0.0043 |
| More than 1 year | 41 (55.4) | 18 (64.3) | 13 (43.3) | 5 (62.5) | 5 (62.5) |  |
| Less than 1 year | 33 (44.6) | 10 (35.7) | 17 (56.7) | 3 (37.5) | 3 (37.5) |  |
| Median time in years (IQR), if more than one year (n=41) | 5 (3-10) | 8.5 (3-20) | 5 (2-8) | 4 (2-8) | 5 (2-6) | 0.2685 |

\* One participant declined to answer the duration of incarceration question

Over half of participants (53.6%) reported experiencing incarceration, varying significantly by site, from 36% at Site D to 70% at Site A. Nearly all incarceration (90.7%) was reported happening more than one year ago, and most (55.4%) reported incarceration lasting more than one year. Of the 41 participants (29.3% overall) reporting incarceration lasting more than one year, the median (IQR) duration of incarceration was 5 (3-10) years; duration was highest at Site A: 8.5 (3-20) years.

Overall, 60 (42.9%) residents reported experiencing both homelessness and incarceration.

### Resident preferences for future services

Figure 1. displays resident preferences for future services, indicated by those who said they were “somewhat interested” or “very interested” in the service. The top five services picked by residents were dental services (92.9%), vision care (92.9%), a facility that accepts TennCare or Medicaid (91.4%), provides adult primary care (91.4%) and serves as a food pantry (90%). TennCare is Tennessee’s state-run Medicaid program.

Looking only at the “very interested” responses, vision care (77.9%) remained the top preferred service, followed closely by dental services (76.4%) and a facility that accepts TennCare or Medicaid (76.4%).

**Figure 1.**
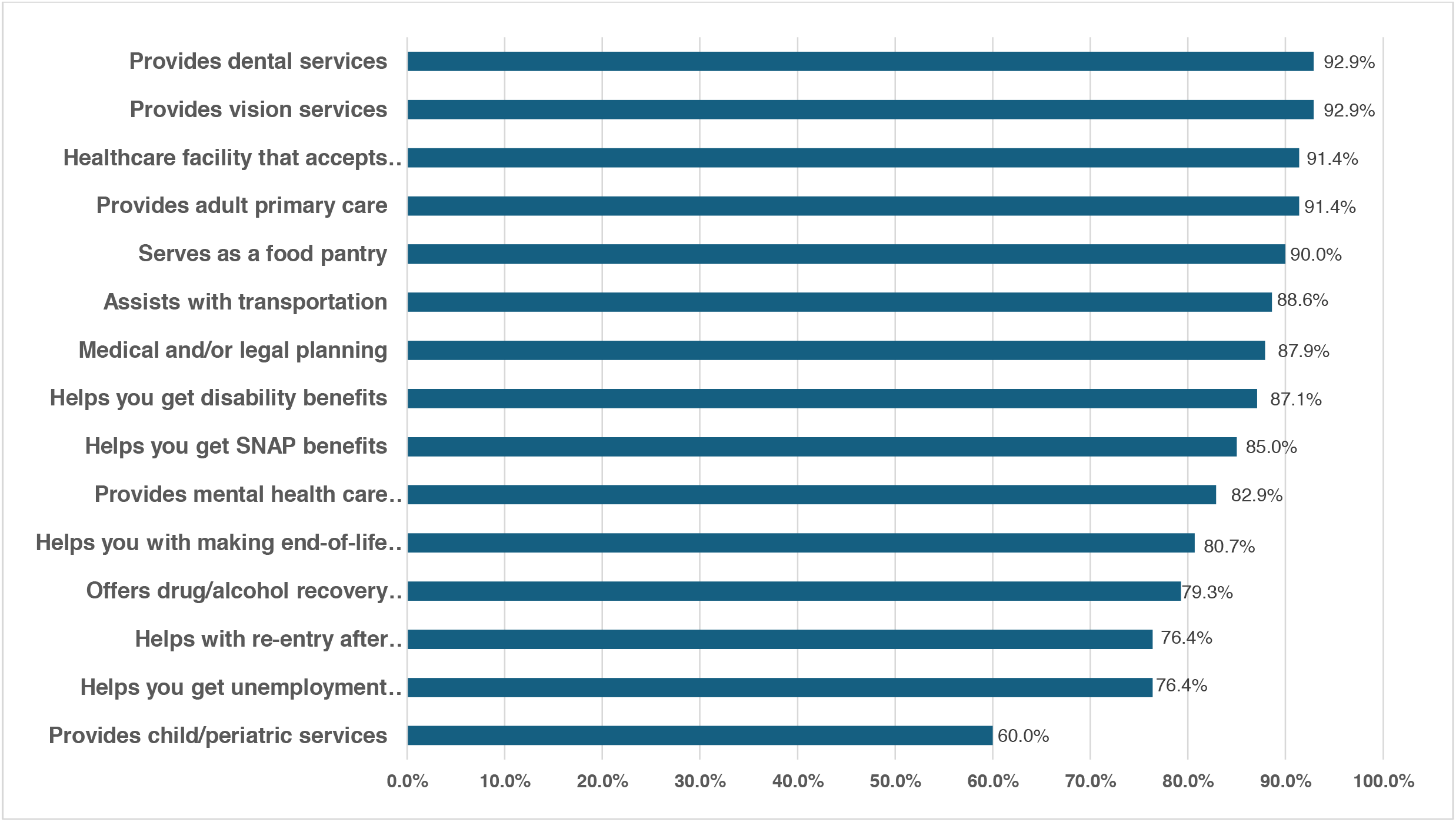
Resident interest in future health and social services provided at a subsidized housing facility in Nashville, TN, 2023-2025 (n=140).

## Discussion

This study explored the demographic, economic, and experiential characteristics of residents living in four nonprofit-owned, subsidized housing facilities in Nashville, Tennessee. Several findings highlight the complex vulnerabilities of this population. Most residents were Black and older adults, and many had low educational attainment and low incomes, with over 80% reporting annual household income $20,000 or less. This profile reflects the economic precarity often seen in subsidized housing communities.^24,25^ These social and economic markers have been strongly associated with poor health outcomes, reduced access to care, and increased exposure to psychosocial stressors.

A key distinction of this study is its setting within nonprofit-managed housing, which differs from publicly owned housing in both resident composition and service structure. Most residents were unpartnered older adults living alone, characteristics that increase the risk of chronic social isolation. Prior studies have emphasized the detrimental health impacts of loneliness and disconnection, particularly among older adults and individuals with a history of trauma or housing instability.^7,26^ More than 60% of participants lacked access to a household car, and 46% received Social Security disability benefits, suggesting a high prevalence of disability. These barriers may further restrict mobility, reduce access to services, and exacerbate isolation. These findings align with broader evidence that social support and community engagement are powerful mechanisms for psychosocial recovery and stability.^26^

A particularly concerning finding was the high prevalence of both lifetime homelessness and incarceration: 42.9% of residents reported experiencing both. Subsidized housing often represents a critical first step in exiting homelessness^2^ and reentering society following incarceration.^27^ However, continued housing instability, especially among individuals with criminal records or prior evictions, remains a persistent barrier to well-being.^28^

Permanent supportive housing (PSH) emerged as an alternative to “treatment first” models, which required people experiencing homelessness to demonstrate sobriety or treatment adherence before receiving permanent housing.^29,30^ PSH programs that follow a Housing First approach combine permanent housing with voluntary health, mental health, and substance use services, and have improved housing stability and reduced acute care use among people with disabilities experiencing homelessness.^29,30^ Co-located, low-barrier substance use treatment has also shown promise for engaging residents of housing programs in care.^31^ Although our participants were already housed, their high prevalence of past homelessness, incarceration, and disability suggests they share many of the needs these models are designed to address.

Participants overwhelmingly prioritized dental and vision care, primary care that accepts Medicaid/TennCare, and food access programs when asked to identify preferred on-site services. These results are consistent with other housing studies where oral health, vision, accessible primary care, and nutrition emerge as top unmet needs.^32,33^ A similar study found that residents identified accessible healthcare services, aging problems, and space for physical activity to be the top three community concerns.^32^ These resident-prioritized services are critical not only for individual health but also for maintaining optimal physical functioning and housing stability. One study site currently partners with a local food pantry to distribute fresh and shelf-stable items monthly, an example of community-based coordination that could be scaled or expanded. Effective implementation of these resident-identified priorities requires partnerships beyond the housing agency, with integrated care models that bridge housing, health, and social systems.^31^

Several methodological strengths enhance the interpretability of this study. Randomized selection of units reduced investigator selection bias, although nonresponse may limit representativeness. The inclusion of bilingual materials and staff increased accessibility. Nevertheless, linguistic diversity beyond English and Spanish may have been underrepresented, and nationwide immigration enforcement activities during the study period may have deterred participation, especially among undocumented residents.

Additionally, the university payment processing service mandated a different payment system for non-US citizens. Unlike their US counterparts, these individuals could not receive physical gift cards; instead, they were required to register as suppliers with the university, and taxes would be withheld from their payments. While these policies are designed to ensure legal compliance, they undoubtedly affect researchers’ efforts to obtain a representative sample of participants and promote equity in research participation. Additionally, while the study offered evening and weekend data collection to accommodate varied work schedules, we may have missed individuals who were unable to participate due to work obligations. Finally, the study’s cross-sectional design limits interpretations of causality and change over time.

By illuminating the demographic profile, structural barriers, and service preferences of subsidized housing residents in the Southeastern United States, this study contributes to an important yet understudied dimension of housing research. Findings underscore the importance of interdisciplinary and population-specific care models embedded within housing environments.^34^ Approaches that build social connections, integrate behavioral health care, and address systemic inequities may help interrupt cycles of marginalization and support residents’ reintegration into community life. Housing providers, policymakers, researchers and health systems should collaborate to transform housing into a true platform for health and equity.

## Public Health Implications

This study provides critical insights into the lived experiences, needs, and service preferences of residents living in nonprofit-owned, subsidized housing in the Southeastern United States, an underexamined yet increasingly vulnerable region. The findings reveal a population marked by structural disadvantage, with high rates of racial and economic marginalization, social isolation, and histories of homelessness and incarceration. Despite the stability that subsidized housing provides, the persistence of unmet health and social needs underscores that housing alone is insufficient to improve population health or promote equity.

Participants expressed strong interest in on-site services, particularly dental, vision, primary care, and food access, which are essential but often inaccessible through traditional service delivery systems. Their preferences reinforce the need for co-located, low-barrier, and trauma-informed interventions that align with residents’ lived realities.

These data support a growing body of evidence calling for interdisciplinary models of care embedded within housing environments, particularly those that center community engagement, address social determinants of health, and build pathways to long-term wellness.^34^

By centering resident voices in program design, this study strengthens the case for reimagining subsidized housing as not merely a form of shelter, but as a platform for integrated, community-driven solutions to health inequities. Future research should prioritize longitudinal evaluation of service models in housing contexts, and policy leaders must invest in scalable strategies that bridge housing and health. Sustainable solutions will depend on cross-sector collaboration that views housing not as a siloed intervention, but as foundational infrastructure for equitable health systems.^35^ The findings from studies like this should be discussed in collaboration with housing providers to ensure they are practical and actionable, ultimately leading to meaningful steps following the research.

## Data Availability

De-identified data produced in the present study are available upon reasonable request to the authors

## Acknowledgements

The study team is deeply grateful to the residents who participated and shared their experiences. We thank the staff of Urban Housing Solutions, especially Brent Elrod, Tiffany Davis, Amber Freeman, Tyneshia Jackson, and Jamie Wesley. We also appreciate the contributions of Dr. Mary Dietrich for randomization and early statistical support.

